# The First General Consent Implementation in Swiss Traditional Chinese Medicine Practices. A Prospective One-Year Study

**DOI:** 10.1101/2024.08.18.24312166

**Authors:** Xiaying Wang, Xiaoying Lv, Bingjun Chen, Saroj Pradhan, Ralf Bauder, Yiming Li, Michael Furian

**Affiliations:** Research Department, Swiss University of Traditional Chinese Medicine (SWISS TCM UNI), Bad Zurzach, Switzerland; Institute Of Basic Research In Clinical Medicine, China Academy Of Chinese Medical Sciences, Beijing, China; Wuxi Traditional Chinese Medicine Hospital, Jiangsu, China

## Abstract

**Background:** Traditional Chinese Medicine (TCM) encompasses a wide range of treatments focused on diagnosing and managing illnesses, with increasing adoption in Western countries. TCM is often applied in isolated practices, therefore, rigorous research and real-world data collection remain challenging. The implementation of the General Consent (GC) facilitates this research, therefore, the aim was to investigate the acceptance rate of the GC during the first year of its implementation in TCM practices.

**Methods:** This prospective cohort study was conducted from 1^st^ January to 31^st^ December 2023 in five TCM practices located in Bad Zurzach, Baden, Lenzburg, Wil, and Zug in Switzerland. GC, together with other registration forms, were sent to patients prior to their appointments, collected during their first visit, and recorded by clinic secretaries. Logistic regression analysis was performed to investigate demographic factors influencing GC acceptance, considering variables such as age, sex, and practice location.

**Results:** The study recorded 1,095 patients who sought TCM treatments in 2023, of which 73.6% returned a valid GC document. Overall, the GC acceptance rate was 611/1,095 (55.8%); of those returning the GC, the acceptance rate was 611/806 (75.8%). The median[IQR] age of patients was 52[37,64] years and female patients were twice as likely to seek TCM treatments compared to male patients. Logistic regression analysis, odds ratio (95%CI), revealed no difference in GC acceptance rate with older age: 1.015 (0.996 to 1.034), p=0.115; female sex: 1.847 (0.588 to 5.804), p=0.294 and age*female sex: 0.983 (0.962 to 1.004), p=0.119. Significant differences in GC acceptance rates were observed across the five TCM practices.

**Conclusion:** The GC implementation in TCM practices was feasible, and the GC was well accepted by patients, independent of sex and age. The observed practice-related differences in GC acceptance require further investigation. More TCM practices should implement the GC to enable practice-based TCM research.

## Background

Traditional Chinese Medicine (TCM) encompasses a wide array of treatment methods focused on diagnosing and managing illnesses. Its adoption has markedly increased in Western countries, further supported by the World Health Organization’s inclusion of TCM diagnostic patterns in the latest update of the International Classification of Diseases code (version 11, accepted in 2019), which serves as the universal benchmark for health diagnostics [1]. In Switzerland, a 2017 survey by the Swiss Federal Statistical Office revealed that 8.5% of respondents (1,592 out of 18,832) had used TCM or acupuncture in the past 12 months [2]. Assuming that this survey is representative of Switzerland, a total of 598,655 of 7,043,002 (corresponding to 8.5%) Swiss inhabitants might have used TCM in 2022. Given the widespread TCM presence and its possible significant impact on contemporary healthcare practices and clinical routines, it is crucial to rigorously evaluate and conduct research on the efficacy of TCM therapies in various diseases [3].

To conduct clinical research ethically and effectively, obtaining informed consent (IC) is recognized as a foundational requirement, mandated by national and international standards like the International Conference on Harmonization -Good Clinical Practice (ICH-GCP) [4]. It ensures participants’ autonomy by fully informing them about a specific study’s objectives, methods, potential benefits, and risks, securing their voluntary agreement [5]. On the other hand, General Consent (GC) is a flexible alternative addressing some of the impracticability of informed consent [6]. It permits the use of participants’ data or biological samples obtained during the clinical routine for future yet-to-be-defined research projects by streamlining research processes, enhancing the potential for scientific breakthroughs, and upholding ethical standards through voluntary, informed participation [7], [8].

The widespread adoption of the GC in various research fields and institutions highlights its advantages [9]. In conventional medicine and hospitals, GC has been a standard element of the admissions process for all in-and outpatients [8]. Yet, its utilization within TCM research and practices is lacking compared to the usage of ICs [10]. The varied modalities of TCM, such as herbal medicine and acupuncture, present distinct obstacles for clinical studies due to the personalized nature of its treatments [11]. Therefore, adopting GC in TCM research will play a crucial role in facilitating its progress and addressing the unique challenges it faces.

In this study, we pioneer the introduction of GC within the context of TCM research, aiming to reconcile TCM’s traditional approaches with modern ethical standards. This initiative seeks to streamline the research process in TCM, facilitating wider research activities and deeper investigation into real-world TCM’s effects and mechanisms while maintaining ethical integrity through informed, voluntary participation.

Therefore, the purpose of this study was to investigate the GC acceptance rate and demographic factors influencing a patient’s GC choice during the first year of GC implementation in five TCM practices in Switzerland.

## Methods

This prospective study was conducted from the 1^st^ of January to the 31^st^ of December 2023 at five TCM practices of the TCM Ming Dao AG situated in Bad Zurzach, Baden, Lenzburg, Wil, and Zug in Switzerland. We used completely anonymous data, which does not require informed consent from participants or approval from the ethics committee according to local law [12]. Our study adhered to STROBE guidelines (STrengthening the Reporting of OBservational studies in Epidemiology) [13]. The GC has been adapted from the Swissethics template [14] and was approved by the Ethics Committee of Northwest-Central Switzerland (No. AO_2023-00017). The GC was available in German and English language.

## Study population

This study involved patients who sought TCM treatments at any of the five outpatient practices during the year 2023. There were no specific age, sex, or literacy requirements for inclusion and no sample size estimation was performed. Eligible participants were required to be conscious adults (in the case of patients aged under 18, their legal guardians) capable of understanding the nature and implications of the German or English GC version.

## Data collection and data management

In case a patient contacted the TCM practice secretary and arranged a TCM treatment, the GC and other administrative documents were sent by mail to the patient. During the patient’s admission and the first visit, the GC was discussed, if required, and the completed and signed GC was collected by the secretary’s office. As part of the daily routine, the secretary’s offices entered the GC information into an internal file containing all new patients seeking TCM therapy. The decision of a patient to accept or reject the GC was not communicated to the TCM providers and either decision did not affect the anticipated TCM treatment.

## TCM Practices

The five TCM practices differed in terms of organizational structures. The practice located in Bad Zurzach had three people working in the secretary office; Baden and Lenzburg had one secretary person while the TCM providers in Wil and Zug had no secretary support. In the five TCM practices, a total of 11 TCM providers were actively treating patients.

## Statistical analysis

We analyzed the observations through descriptive statistics for patients agreeing, declining, or not issuing and incorrectly completing GCs. A GC was not issued or invalid (GC status = not issued/invalid) if the forms were not handed out to the patients or not returned to the secretary or if the document contained missing information, i.e., missing signature of the patient. Results are presented in median (interquartile ranges) or absolute numbers and percentages. GC acceptance was stratified and analyzed over sex and different age groups.

We performed two logistic multivariable regression analyses to investigate whether demographic characteristics, the time since GC implementation, or the TCM practices were predictors for the GC issues and the GC acceptance. In the former, issued GC, i.e., valid GC document returned to the secretary, and not issued/invalid GC was the dependent binary variable. In the latter, GC acceptance (Yes, No, excluding the not issued/invalid GC) was the dependent binary variable. Age, sex (men, women), interaction between age and sex, and TCM practices (Bad Zurzach, Baden, Lenzburg, Wil, Zug) were independent predictors. The interaction effects between sex and age were also analyzed to evaluate how the impact of age on the GC agreement might differ depending on sex. The time since implementation was calculated as the difference in months of patients’ first visit’s date from the 1^st^ of January 2023 when the GC was first introduced.

Statistical analyses were performed with STATA v15 and RStudio version 4.3.3. A p-value below 0.05 was considered to reflect statistical significance.

## Results

### Population overview

As reported in Figure 1, a total of 1,095 patients visited the five practices from January to December 2023, with Bad Zurzach receiving the most of them (50.8%), followed by Baden (22.6%), Lenzburg (16.3%), Zug (7.9%), and Wil (2.5%). Out of the total amount of records, in 289 of 1,095 (26.4%) of the cases, the GC was either not issued (15 of 289 (5.2%) cases due to language issues, other reasons remain unknown), or the patient did not provide a valid GC document. The remaining 806 of 1,059 (73.6%) signed a GC, of which 611 of 806 (75.8%) agreed, and 195 of 806 (24.2%) declined. In the context of all new patients, a positive GC was obtained from 611 of 1,095 (55.8%) cases (Figure 1).

**Figure 1.**
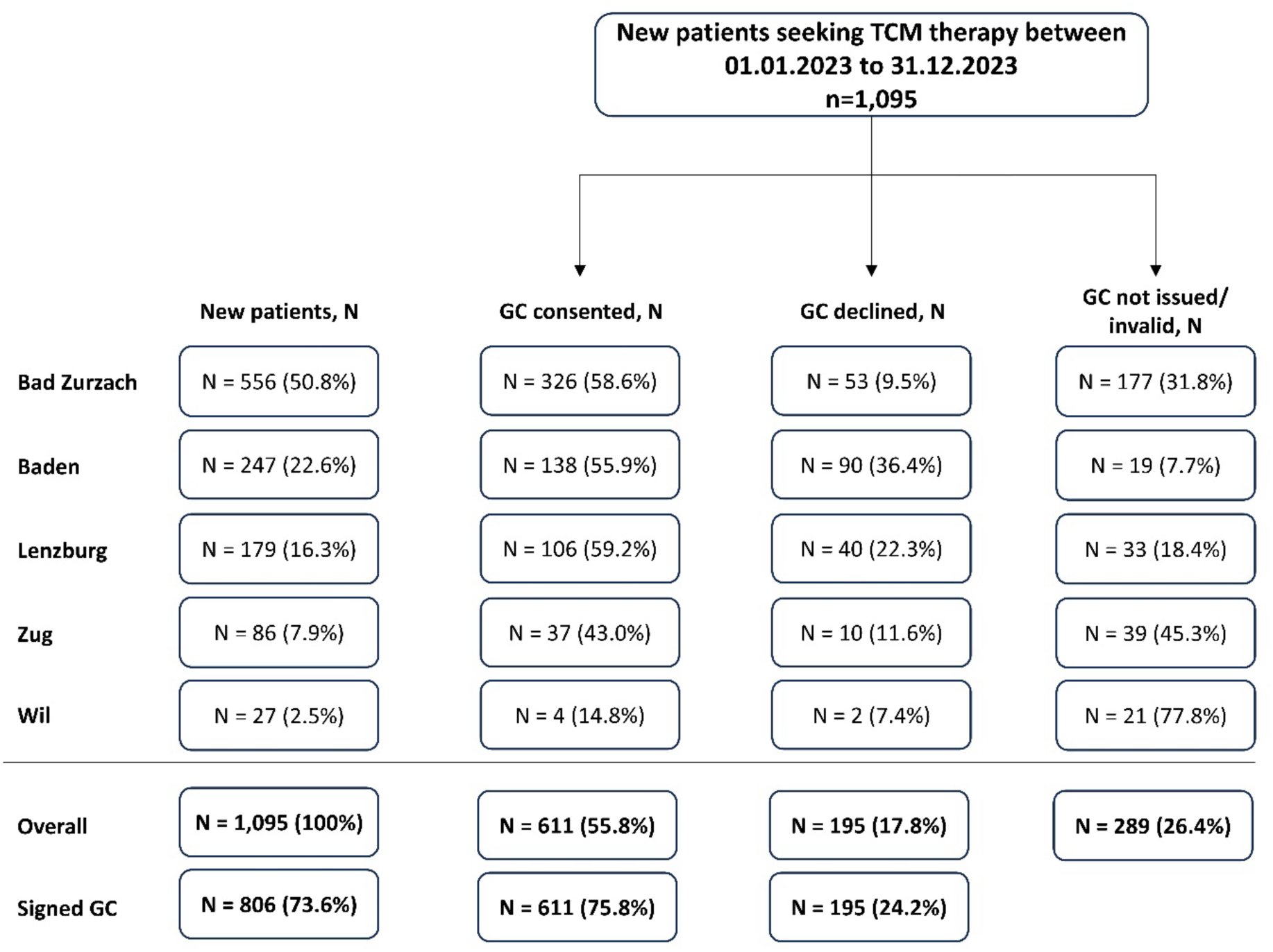
Patient flow chart stratified by TCM practice and by General Consent categorization. TCM, traditional Chinese medicine; GC, General Consent.

### Demographics

Around two-thirds of the new patients seeking TCM treatments in 2023 were female, of which 54.9% gave their consent, and 19.1% declined the GC (Table 1, Figure 2). Similar proportions of GC status were found in male patients (n = 357, 58.0% consented, 15.4% declined), P = NS between sexes.

**Figure 2.**
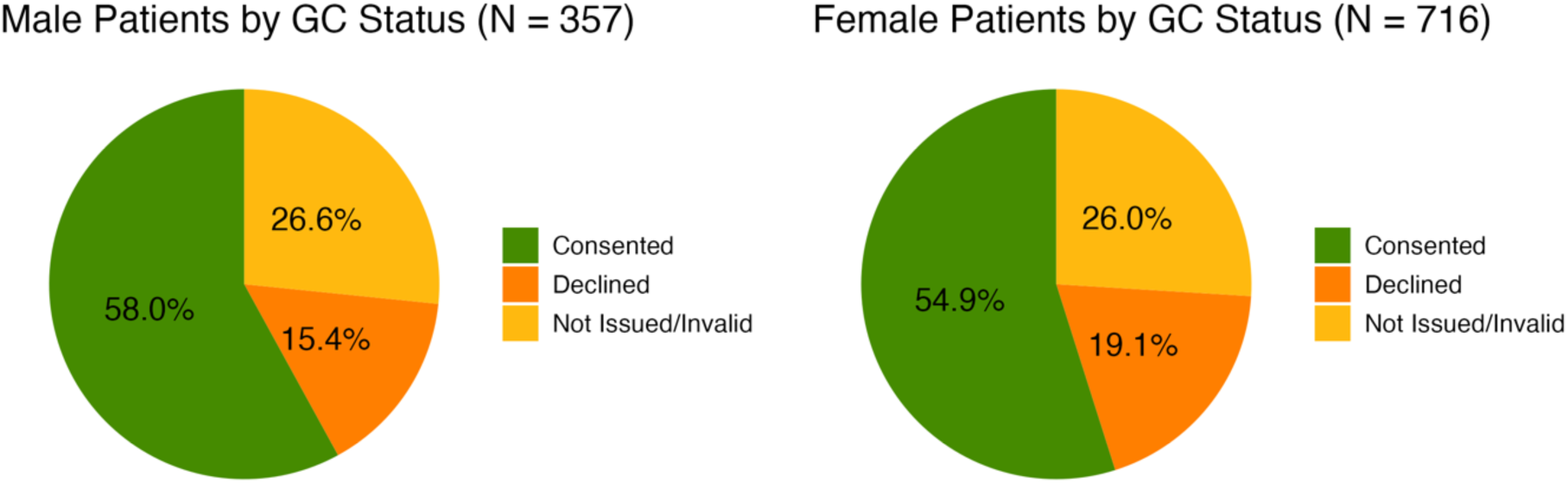
Sex-related General Consent categorization. GC, General Consent.

**Table 1.** Demographic characteristics of new patients seeking TCM therapy in 2023.

| **Variable** | **New patients** | **Consented** | **Declined** | **Not issued/invalid** |
| --- | --- | --- | --- | --- |
| N | 1,095 (100) | 611 (55.8) | 195 (17.8) | 289 (26.4) |
| Sex (n, %) |  |  |  |  |
| Male: | 357 (32.6) | 207 (58.0)* | 55 (15.4) | 95 (26.6) |
| Female: | 716 (66.4) | 393 (54.9)* | 137 (19.1) | 186 (26.0) |
| Unknown: | 22 (2.0) | 11 (50%) | 3 (13.6) | 8 (36.4) |
| Age, years | 52 [37, 64] | 52 [38, 65] | 48 [35.5, 61] | 53 [37, 64] |

Values are presented in numbers (proportions) or median [interquartile range]. \*P<0.05 between the proportion of patients consenting compared to declining within sex.

The median [interquartile range] age of all new patients was 52 [37, 64] years old. The relationship between age and GC acceptance rate is illustrated in proportion and in absolute years in Figure 3 and Figure 4. However, the logistic regression analysis revealed that neither sex nor age played a role in the GC acceptance rate (Table 2 and Table 3).

**Figure 3.**
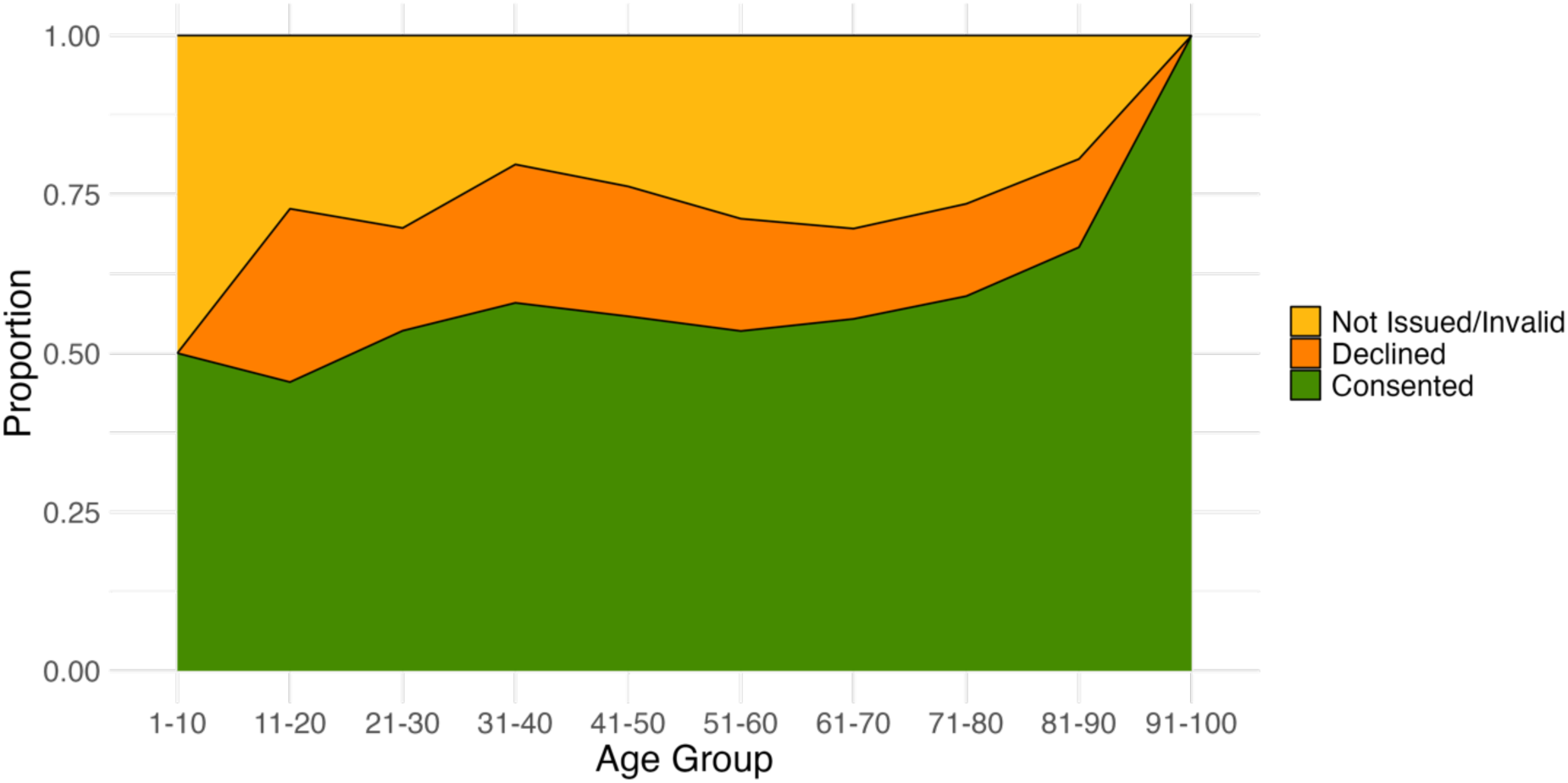
Proportional distribution of the General Consent status divided into age decades.

**Figure 4.**
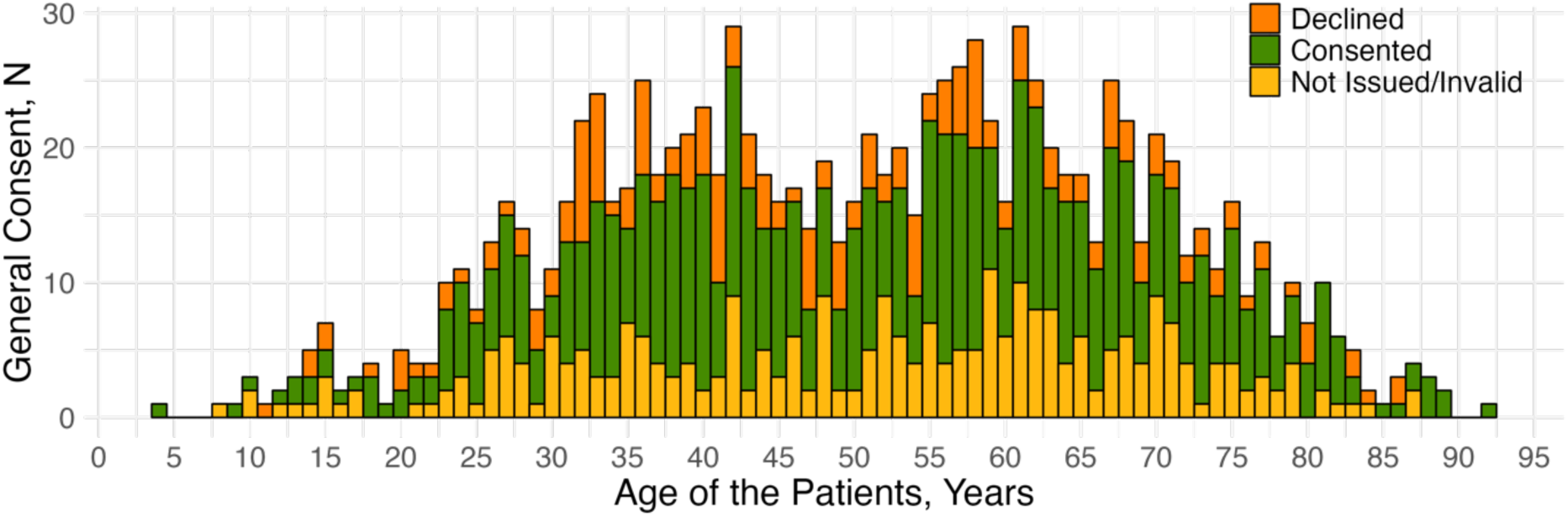
Distribution of General Consent status in all ages.

**Table 2.** Logistic regression – General Consent implementation: not issued/invalid vs. issued.

| **Variable** | **Odds ratio** | **95% confidence interval** | **p-value** |
| --- | --- | --- | --- |
| Age, years | 1.008 | 0.945 to 1.076 | 0.803 |
| Female sex (*reference: male*) | 2.209 | 0.130 to 37.546 | 0.584 |
| Female * Age | 0.993 | 0.930 to 1.061 | 0.841 |
| Months since GC implementation | 1.073 | 1.027 to 1.120 | 0.001* |
| **TCM practice:** |  |  |  |
| *reference (Bad Zurzach)* |  |  |  |
| Baden | 5.898 | 3.554 to 9.786 | <0.001* |
| Lenzburg | 2.308 | 1.486 to 3.586 | <0.001* |
| Zug | 0.633 | 0.394 to 1.018 | 0.060 |
| Wil | 0.142 | 0.056 to 0.359 | <0.001* |

N = 1,094. \*P<0.05 compared to the reference

**Table 3.** Logistic regression – General Consent status (excluding not issued/invalid, i.e., analyzing the issued ones): declined vs. consented.

| Variable | Odds ratio | 95% confidence interval | p-value |
| --- | --- | --- | --- |
| Age, years | 1.015 | 0.996 to 1.034 | 0.115 |
| Female sex (*reference: male*) | 1.851 | 0.589 to 5.822 | 0.292 |
| Female * Age | 0.983 | 0.962 to 1.005 | 0.120 |
| Months since GC implementation | 1.046 | 0.992 to 1.103 | 0.096 |
| **TCM practice:** |  |  |  |
| *reference (Bad Zurzach)* |  |  |  |
| Baden | 0.246 | 0.164 to 0.369 | <0.001* |
| Lenzburg | 0.431 | 0.266 to 0.700 | 0.001* |
| Zug | 0.584 | 0.272 to 1.253 | 0.167 |
| Wil | 0.299 | 0.053 to 1.697 | 0.173 |

N = 792. \*P<0.05 compared to the reference

### Impact of the TCM practices

When observing the GC status within each TCM practice (Figure 1), Bad Zurzach presented 58.6% of newly registered patients agreeing to the GC, and 31.8% of them did not receive it or did not provide a valid GC document. In Baden, Lenzburg, and Zug, the GC agreement rate was 55.9%, 59.2%, and 43.0%, respectively (P = NS compared to Bad Zurzach). In Wil, the GC acceptance rate was 14.8% (P<0.05 compared to Bad Zurzach) and the percentage of not issued/invalid GC was the highest, i.e., 77.8% (P<0.05 compared to all other practices). This is confirmed by the multivariable logistic regression analysis on GC issues, where not issued/invalid GC is significantly more likely to happen in Wil compared to Bad Zurzach (P<0.001), whereas it is significantly more likely to obtain a returned, valid GC in Baden and Lenzburg (P<0.001) (Table 2).

In the subgroup of patients providing a valid GC statement, i.e. the patients consenting or rejecting the GC, excluding the not issued and invalid GC, the multivariable logistic regression analysis revealed that patients in Baden and in Lenzburg were significantly less likely to accept the GC compared to patients in Bad Zurzach (Table 3). This was likely due to the much higher rate of “not issued and invalid” GCs in Bad Zurzach compared to Baden and Lenzburg.

### Time since GC implementation

We analyzed whether the implementation of the GC became more effective with time by assessing the return of a valid GC document, whether it was agreed upon or declined, compared to not issued and invalid GC documents (Table 2). The results show that the issuing of GCs significantly improved over time (odds ratio, 95%CI, 1.0.73, 1.027 to 1.120, P=0.001). However, the time since implementation did not have any effect on the GC acceptance rate (odds ratio, 95%CI, 1.046, 0.992 to 1.103, P=0.096) (Table 3).

## Discussion

The current study provides the first insights into the implementation and acceptance rate of GC in TCM practices. We found that the implementation of the GC in TCM practices is feasible and that the GC acceptance rate varies across practices but remains relatively high. Importantly, the GC acceptance rate was independent of age and sex, suggesting that future TCM research, depending on the GC acceptance rate, will presumably not be biased towards age and sex. These findings might encourage other TCM practices to implement the GC, facilitating future practice-based TCM research using patient’s data obtained in daily clinical routine.

Despite numerous studies on IC and ethical considerations [10], [15], [16], [17], [18], there has been no work on GC in TCM research. Our results showed a considerable acceptance rate (55.8%) of GC among all incoming new patients. This is 17% higher compared to a study on GC implementation at the University Hospital Zurich (USZ) [8] (55.8% vs. 38.8%) (P<0.001, two-sample test of proportions), thanks to the much lower rate of not issued or invalid GCs (26.4% vs. 51.3%) (P<0.001, two sample test of proportions). However, out of those who returned a valid GC document, 75.8% consented to participate. This is significantly lower than the 79.7% of the returned and agreed GCs at the USZ [8] (P<0.05, two sample test of proportions). Although, lower, the generally high acceptance rates suggest that patients are open to research participation beyond conventional medical frameworks and centralized hospital settings and show a similar willingness to provide their patient data to complementary medicine research in a decentralized setting like TCM practices.

The TCM practices that had the least amount of not issued/invalid GC was the one in Baden (7.7%), whereas the practice in Wil experienced the highest rate (77.8%, P<0.001 compared to Bad Zurzach) followed by Zug (45.3%, P=NS compared to Bad Zurzach). The underlying reason, remains not well understood but could be related to differences in the patient population, time constrains or due to structural differences in the TCM practices or other unknown reasons. For example, Zug and Wil had no secretary support, therefore, the TCM providers might have been under time constrains and might have prioritized the medical treatment over the GC implementation. Whereas Baden and Lenzburg showed significantly lower not issued/invalid GCs compared to Bad Zurzach (P<0.001). The reason could be the simpler organizational structure in Baden and Lenzburg, where there were each one secretary person and one TCM practitioner compared to the three secretary people and 4 – 5 practitioners in Bad Zurzach. The analysis on the time since implementation showed that the chance that a GC was issued increased on average by 7.3% / month over the time period of one year (P<0.001, Table 2). This might be due to the staff’s familiarization and improvement with handling the GC process.

During the year 2023, female patients seeking TCM treatments were twice as many as male patients. This greater presence of the female population in TCM usage is consistent with findings from a study conducted in Taiwan, where the one-year prevalence of TCM use was 31.8% for women and 22.4% for men, out of 14,064 eligible participants [19]. Although specific studies on TCM usage in Europe are lacking, studies have shown that women generally seek complementary and alternative medicine (CAM) treatments more frequently than men. More specifically, a study from Norway indicated that 42% of those reporting CAM usage were female compared to 24% male [20]. Similarly, a broader and more recent study in Europe showed that 21.5% of female and 13.9% of male participants reported usage of CAM [21].

Despite the higher proportion of female patients seeking TCM therapies, the GC acceptance rate remained independent of sex and age (Table 3). This finding seems to differ from the findings of the GC study conducted at the University Hospital Zurich, where age was positively and female sex negatively affecting the GC acceptance rate. These differences highlight that GC acceptance rates might depend on the settings and are not generalizable across different fields of medicine. Nevertheless, the inexistent correlation of sex and age with the GC acceptance rate presents a promising finding, suggesting that future TCM research conducted on the basis of GCs will likely not be biased towards sex or age.

One of the strengths of this study is its pioneering nature in implementing GC in TCM practices, successfully bridging traditional medical practices with modern ethical standards. It paves the way for practices-based TCM research in the real-world setting. Moreover, the successful implementation provides a framework for other practices and institutions to adopt similar practices. However, the study faces several limitations that affect its overall robustness and generalizability. The focus on five TCM practices in the German-speaking part of Switzerland limits the ability to generalize the findings to a broader population or different geographic and cultural contexts. The logistic regression model primarily considered age, sex, and practice location, excluding other potentially influential factors like socioeconomic status, education level, and specific health conditions.

## Conclusion

This prospective study describes the first implementation of General Consent (GC) in Traditional Chinese Medicine (TCM) practices in Switzerland. Findings revealed that the GC implementation was feasible in five TCM practices and that patients, independent of age and sex, showed a high GC acceptance rate. These findings might encourage other practices to implement the GC to facilitate practice-based, real-world research in TCM and beyond. Establishing a network of TCM practices willing to participate in research, along with nationwide implementation of the GC and standardized entry and exit forms, will enable novel and highly warranted research in the field of TCM.

## Availability of data and materials section

The datasets generated and/or analyzed during the current study are not publicly available due to the protection of patient data but anonymized data are available from the corresponding author upon reasonable request.

## Supporting information

STROBE Statement

## Data Availability

All data produced in the present study are available upon reasonable request to the authors

## Authors’ contributions

XW completed the statistical analysis, designed and made the graphs/figures, and wrote the first draft of the manuscript. XL and BC structured and acquired the data. RB and SP assisted with the GC implementation and acquisition of the data. MF supervised the planning, conduct, and completion of the project, and assisted XW with statistical analysis. MF and YL conceptualized and designed the project. All authors critically reviewed and approved the final manuscript.

## References

[1] “ICD-11.” Accessed: Mar. 21, 2024. [Online]. Available: https://icd.who.int/en

[2] D. Meier-Girard, E. Lüthi, P. Y. Rodondi, and U. Wolf, “Prevalence, specific and non-specific determinants of complementary medicine use in Switzerland: Data from the 2017 Swiss Health Survey,” PLoS One, vol. 17, no. 9, p. e0274334, Sep. 2022, doi: 10.1371/JOURNAL.PONE.0274334.

[3] M. Eigenschink, L. Dearing, T. E. Dablander, J. Maier, and H. H. Sitte, “A critical examination of the main premises of Traditional Chinese Medicine,” Wiener Klinische Wochenschrift, vol. 132, no. 9–10. Springer Medizin, pp. 260–273, May 01, 2020. doi: 10.1007/s00508-020-01625-w.

[4] A. Vijayananthan and O. Nawawi, “The importance of Good Clinical Practice guidelines and its role in clinical trials,” Biomedical Imaging and Intervention Journal, vol. 4, no. 1. Jan. 2008. doi: 10.2349/biij.4.1.e5.

[5] European Medicines Agency, “Guideline for good clinical practice E6(R2) [Internet]. European Medicines Agency.” Accessed: Mar. 22, 2024. [Online]. Available: https://www.ema.europa.eu/en/documents/scientific-guideline/ich-e-6-r2-guideline-good-clinical-practice-step-5_en.pdf

[6] S. J. M. Laurijssen et al., “When is it impractical to ask informed consent? A systematic review,” Clinical Trials, vol. 19, no. 5, pp. 545–560, Oct. 2022, doi: 10.1177/17407745221103567/ASSET/IMAGES/LARGE/10.1177_17407745221103567-FIG2.JPEG.

[7] M. G. Hansson, J. Dillner, C. R. Bartram, J. A. Carlson, and G. Helgesson, “Should donors be allowed to give broad consent to future biobank research?,” Lancet Oncology, vol. 7, no. 3, pp. 266–269, Mar. 2006, doi: 10.1016/S1470-2045(06)70618-0.

[8] A. Griessbach, A. Bauer, F. J. Lebet, and R. Grossmann, “The concept of General Consent in Switzerland and the implementation at the University Hospital Zurich, a cross-sectional study,” Swiss Med Wkly, vol. 152, no. 15–16, Apr. 2022, doi: 10.4414/smw.2022.w30159.

[9] R. B. Mikkelsen, M. Gjerris, G. Waldemar, and P. Sandøe, “Broad consent for biobanks is best-provided it is also deep,” BMC Med Ethics, vol. 20, no. 1, pp. 1–12, Oct. 2019, doi: 10.1186/S12910-019-0414-6/PEER-REVIEW.

[10] H. Zhao, J. Zhang, F. Yang, and L. Tan, “Improve the ethical review of clinical trials on traditional medicine: A cross-sectional study of clinical trial registration, ethical review, and informed consent in clinical trials of Traditional Chinese Medicine,” Medicine, vol. 97, no. 47, Nov. 2018, doi: 10.1097/MD.0000000000013062.

[11] J. Zhang and Z. M. Zhang, “The Challenges of Ethical Review in Clinical Research of Traditional Chinese Medicine,” Evid Based Complement Alternat Med, vol. 2021, 2021, doi: 10.1155/2021/6754985.

[12] “Ethikkommission Nordwest-und Zentralschweiz -EKNZ.” Accessed: Mar. 22, 2024. [Online]. Available: https://www.eknz.ch/

[13] J. P. Vandenbroucke et al., “Strengthening the Reporting of Observational Studies in Epidemiology (STROBE): explanation and elaboration,” Int J Surg, vol. 12, no. 12, pp. 1500–1524, Dec. 2014, doi: 10.1016/J.IJSU.2014.07.014.

[14] swissethics - Swiss Association of Research Ethics Committees, “Generalkonsent,” swissethics. Accessed: Apr. 02, 2024. [Online]. Available: https://swissethics.ch/documents/generalkonsent

[15] X. Liu et al., “Informed consent in cancer clinical drug trials in China: a narrative literature review of the past 20 years,” Trials, vol. 24, no. 1, Dec. 2023, doi: 10.1186/S13063-023-07482-Y.

[16] X. Y. Wang, Z. H. Liang, H. L. Huang, and W. X. Liang, “Principles of ethics review on traditional medicine and the practice of institute review board in China,” Chin J Integr Med, vol. 17, no. 8, pp. 631–634, Aug. 2011, doi: 10.1007/S11655-011-0820-1.

[17] C. A. Smith, R. Priest, B. Carmady, S. Bourchier, and A. Bensoussan, “The Ethics of Traditional Chinese and Western Herbal Medicine Research: Views of Researchers and Human Ethics Committees in Australia,” Evid Based Complement Alternat Med, vol. 2011, 2011, doi: 10.1155/2011/256915.

[18] K. Chatfield, B. Salehi, J. Sharifi-Rad, and L. Afshar, “Applying an Ethical Framework to Herbal Medicine,” Evid Based Complement Alternat Med, vol. 2018, Jan. 2018, doi: 10.1155/2018/1903629.

[19] C. C. Shih, C. C. Liao, Y. C. Su, C. C. Tsai, and J. G. Lin, “Gender Differences in Traditional Chinese Medicine Use among Adults in Taiwan,” PLoS One, vol. 7, no. 4, p. e32540, Apr. 2012, doi: 10.1371/JOURNAL.PONE.0032540.

[20] A. E. Kristoffersen, T. Stub, A. Salamonsen, F. Musial, and K. Hamberg, “Gender differences in prevalence and associations for use of CAM in a large population study,” BMC Complement Altern Med, vol. 14, no. 1, pp. 1–9, Dec. 2014, doi: 10.1186/1472-6882-14-463/TABLES/5.

[21] E. L. Fjær, E. R. Landet, C. L. McNamara, and T. A. Eikemo, “The use of complementary and alternative medicine (CAM) in Europe,” BMC Complement Med Ther, vol. 20, no. 1, pp. 1–9, Apr. 2020, doi: 10.1186/S12906-020-02903-W/TABLES/3.

