## Supplementary material for "The First General Consent Implementation in Swiss Traditional Chinese Medicine Practices. A Prospective One-Year Study": STROBE Statement

STROBE Statement—checklist of items that should be included in reports of observational studies

| | Item No. | Recommendation | Page No. | Relevant text from manuscript |
|---|---|---|---|---|
| **Title and abstract** | 1 | (*a*) Indicate the study's design with a commonly used term in the title or the abstract | 1 | "The First General Consent Implementation in Swiss Traditional Chinese Medicine Practices. A Prospective One-Year Study." |
| | | (*b*) Provide in the abstract an informative and balanced summary of what was done and what was found | 2 | **"Summary**<br>**Background**: Traditional Chinese Medicine (TCM) encompasses a wide range of treatments focused on diagnosing and managing illnesses, with increasing adoption in Western countries. TCM is often applied in isolated practices, therefore, rigorous research and real-world data collection remain challenging. The implementation of the General Consent (GC) facilitates this research, therefore, the aim was to investigate the acceptance rate of the GC during the first year of its implementation in TCM practices.<br>**Methods**: This prospective cohort study was conducted from 1st January to 31st December 2023 in five TCM practices located in Bad Zurzach, Baden, Lenzburg, Wil, and Zug in Switzerland. GC, together with other registration forms, were sent to patients prior to their appointments, collected during their first visit, and recorded by clinic secretaries. Logistic regression analysis was performed to investigate demographic factors influencing GC acceptance, considering variables such as age, sex, and practice location.<br>**Results**: The study recorded 1,095 patients who sought TCM treatments in 2023, of which 73.6% returned a valid GC document. Overall, the GC acceptance rate was 611/1,095 (55.8%); of those returning the GC, the acceptance rate was 611/806 (75.8%). The median[IQR] age of patients was 52[37,64] years and female patients were twice as likely to seek TCM treatments compared to male patients. Logistic regression analysis, odds ratio (95%CI), revealed no difference in GC acceptance rate with older age: 1.015 (0.996 to 1.034), p=0.115; female sex: 1.847 (0.588 to 5.804), p=0.294 and age*female sex: 0.983 (0.962 to 1.004), p=0.119. Significant differences in GC acceptance rates were observed across the five TCM practices. |

| | | | | |
|---|---|---|---|---|
| | | | | **Conclusion**: The GC implementation in TCM practices was feasible, and the GC was well accepted by patients, independent of sex and age. The observed practice-related differences in GC acceptance require further investigation. More TCM practices should implement the GC to enable practice-based TCM research.” |
| **Introduction** | | | | |
| Background/rationale | 2 | Explain the scientific background and rationale for the investigation being reported | 3 | “Traditional Chinese Medicine (TCM) encompasses a wide array of treatment methods focused on diagnosing and managing illnesses. […] Therefore, adopting GC in TCM research will play a crucial role in facilitating its progress and addressing the unique challenges it faces.” |
| Objectives | 3 | State specific objectives, including any prespecified hypotheses | 4 | “In this study, we pioneer the introduction of GC within the context of TCM research, aiming to reconcile TCM's traditional approaches with modern ethical standards. […]<br>Therefore, the purpose of this study was to investigate the GC acceptance rate and demographic factors influencing a patient's GC choice during the first year of GC implementation in five TCM practices in Switzerland.” |
| **Methods** | | | | |
| Study design | 4 | Present key elements of study design early in the paper | 4 | **“Methods**<br>This prospective study was conducted from the 1st of January to the 31st of December 2023 at five TCM practices of the TCM Ming Dao AG situated in Bad Zurzach, Baden, Lenzburg, Wil, and Zug in Switzerland. We used completely anonymous data, which does not require informed consent from participants or approval from the ethics committee according to local law [12]. Our study adhered to STROBE guidelines (STrengthening the Reporting of OBservational studies in Epidemiology) [13]. The GC has been adapted from the Swissethics template [14] and was approved by the Ethics Committee of Northwest-Central Switzerland (No. AO_2023-00017). The GC was available in German and English language.” |
| Setting | 5 | Describe the setting, locations, and relevant dates, including periods of recruitment, exposure, follow-up, and data collection | 4, 5 | **“Study population**<br>This study involved patients who sought TCM treatments at any of the five outpatient practices during the year 2023. [..] Eligible participants were required to be conscious |

| | | | | |
|---|---|---|---|---|
| | | | | adults (in the case of patients aged under 18, their legal guardians) capable of understanding the nature and implications of the German or English GC version.<br>**Data collection and data management**<br>In case a patient contacted the TCM practice secretary and arranged a TCM treatment, the GC and other administrative documents were sent by mail to the patient. During the patient's admission and the first visit, the GC was discussed, if required, and the completed and signed GC was collected by the secretary's office. As part of the daily routine, the secretary's offices entered the GC information into an internal file containing all new patients seeking TCM therapy. The decision of a patient to accept or reject the GC was not communicated to the TCM providers and either decision did not affect the anticipated TCM treatment.<br>**TCM Practices**<br>The five TCM practices differed in terms of organizational structures. The practice located in Bad Zurzach had three people working in the secretary office; Baden and Lenzburg had one secretary person while the TCM providers in Wil and Zug had no secretary support. In the five TCM practices, a total of 11 TCM providers were actively treating patients." |
| Participants | 6 | (*a*) *Cohort study*—Give the eligibility criteria, and the sources and methods of selection of participants. Describe methods of follow-up<br>*Case-control study*—Give the eligibility criteria, and the sources and methods of case ascertainment and control selection. Give the rationale for the choice of cases and controls<br>*Cross-sectional study*—Give the eligibility criteria, and the sources and methods of selection of participants | 4 | **"Study population**<br>This study involved patients who sought TCM treatments at any of the five outpatient practices during the year 2023. There were no specific age, sex, or literacy requirements for inclusion and no sample size estimation was performed. Eligible participants were required to be conscious adults (in the case of patients aged under 18, their legal guardians) capable of understanding the nature and implications of the German or English GC version." |
| | | (*b*) *Cohort study*—For matched studies, give matching criteria and number of exposed and | | |

| | | | | |
|---|---|---|---|---|
| | | unexposed<br>*Case-control study*—For matched studies, give matching criteria and the number of controls per case | | |
| Variables | 7 | Clearly define all outcomes, exposures, predictors, potential confounders, and effect modifiers. Give diagnostic criteria, if applicable | 5 | Descriptive statistics for demographic characteristics for patients agreeing, declining, or not issuing and incorrectly completing GCs. GC acceptance was stratified and analyzed over sex and different age groups.<br>We performed two logistic multivariable regression analyses with GC issues and GC acceptance as dependent variables and age, sex, interaction between age and sex, and TCM practices as independent predictors. The interaction effects between sex and age were also analyzed to evaluate how the impact of age on the GC agreement might differ depending on sex. The time since implementation was calculated as the difference in months of patients' first visit's date from the 1st of January 2023 when the GC was first introduced. |
| Data sources/ measurement | 8* | For each variable of interest, give sources of data and details of methods of assessment (measurement). Describe comparability of assessment methods if there is more than one group | *5* | The data are collected through GC consent form. |
| Bias | 9 | Describe any efforts to address potential sources of bias | 5 | The logistic regression is an effort to eliminate bias and test the robustness of our conclusions. |
| Study size | 10 | Explain how the study size was arrived at | 5 | The aim was to analyse all patients coming to TCM in the year of 2023. Therefore, no sample size estimation was performed. |

Continued on next page

| | | | | |
|---|---|---|---|---|
| Quantitative variables | 11 | Explain how quantitative variables were handled in the analyses. If applicable, describe which groupings were chosen and why | 5 | Descriptive statistics was used for number of patients, age, sex. Absolute numbers, percentages, and proportions were given. GC acceptance was stratified and analyzed over sex and different age groups to analyze the differences among different groups. The interaction effects between sex and age were also analyzed to evaluate how the impact of age on the GC agreement might differ depending on sex. The time since implementation was also used to analyze if there were significant changes over the year. |
| Statistical methods | 12 | (*a*) Describe all statistical methods, including those used to control for confounding | 5 | Descriptive statistics and logistic multivariable regression analyses. |
| | | (*b*) Describe any methods used to examine subgroups and interactions | 5 | The interaction effects between sex and age were also analyzed to evaluate how the impact of age on the GC agreement might differ depending on sex. It was an independent variable in the logistic regression analyses. |
| | | (*c*) Explain how missing data were addressed | | They were excluded. |
| | | (*d*) *Cohort study*—If applicable, explain how loss to follow-up was addressed<br>*Case-control study*—If applicable, explain how matching of cases and controls was addressed<br>*Cross-sectional study*—If applicable, describe analytical methods taking account of sampling strategy | | There was no follow up needed. |
| | | (*e*) Describe any sensitivity analyses | | NA |
| **Results** | | | | |
| Participants | 13* | (a) Report numbers of individuals at each stage of study—eg numbers potentially eligible, examined for eligibility, confirmed eligible, included in the study, completing follow-up, and analysed | 6, 17 | Results section/Population overview and Figure 1 |
| | | (b) Give reasons for non-participation at each stage | 6, 17 | Not issued or invalid documents are reported in the manuscript. |
| | | (c) Consider use of a flow diagram | 17 | Figure 1 |
| Descriptive data | 14* | (a) Give characteristics of study participants (eg demographic, clinical, social) and information on exposures and potential confounders | 6, 14 | Results section/Demographics and Table 1 |
| | | (b) Indicate number of participants with missing data for each variable of interest | | NA |

| | | | | |
|---|---|---|---|---|
| | | (c) *Cohort study*—Summarise follow-up time (eg, average and total amount) | | NA |
| Outcome data | 15* | *Cohort study*—Report numbers of outcome events or summary measures over time | 6, 17 | Results section/Population overview and Figure 1 |
| | | *Case-control study*—Report numbers in each exposure category, or summary measures of exposure | | *NA* |
| | | *Cross-sectional study*—Report numbers of outcome events or summary measures | | *NA* |
| Main results | 16 | (*a*) Give unadjusted estimates and, if applicable, confounder-adjusted estimates and their precision (eg, 95% confidence interval). Make clear which confounders were adjusted for and why they were included | | 95% confidence interval was reported in all logistic regression analyses. |
| | | (*b*) Report category boundaries when continuous variables were categorized | | NA |
| | | (*c*) If relevant, consider translating estimates of relative risk into absolute risk for a meaningful time period | | NA |

Continued on next page

| | | | | |
|---|---|---|---|---|
| Other analyses | 17 | Report other analyses done—eg analyses of subgroups and interactions, and sensitivity analyses | Figures 2, 3, 4 | Subgroup analyses were done for age and sex |
| **Discussion** | | | | |
| Key results | 18 | Summarise key results with reference to study objectives | 7, 8, 9 | The GC acceptance, issues, number of patients, demographics, and logistic regression analyses were discussed. |
| Limitations | 19 | Discuss limitations of the study, taking into account sources of potential bias or imprecision. Discuss both direction and magnitude of any potential bias | 9 | "However, the study faces several limitations that affect its overall robustness and generalizability. The focus on five TCM practices in the German-speaking part of Switzerland limits the ability to generalize the findings to a broader population or different geographic and cultural contexts. The logistic regression model primarily considered age, sex, and practice location, excluding other potentially influential factors like socioeconomic status, education level, and specific health conditions." |
| Interpretation | 20 | Give a cautious overall interpretation of results considering objectives, limitations, multiplicity of analyses, results from similar studies, and other relevant evidence | 7, 8, 9 | A cautious overall interpretation was given considering objective, limitations, multiple analyses, and comparison with similar studies. |
| Generalisability | 21 | Discuss the generalisability (external validity) of the study results | 9 | "The focus on five TCM practices in the German-speaking part of Switzerland limits the ability to generalize the findings to a broader population or different geographic and cultural contexts." |
| **Other information** | | | | |
| Funding | 22 | Give the source of funding and the role of the funders for the present study and, if applicable, for the original study on which the present article is based | | NA |

*Give information separately for cases and controls in case-control studies and, if applicable, for exposed and unexposed groups in cohort and cross-sectional studies.

**Note:** An Explanation and Elaboration article discusses each checklist item and gives methodological background and published examples of transparent reporting. The STROBE checklist is best used in conjunction with this article (freely available on the Web sites of PLoS Medicine at http://www.plosmedicine.org/, Annals of Internal Medicine at http://www.annals.org/, and Epidemiology at http://www.epidem.com/). Information on the STROBE Initiative is available at www.strobe-statement.org.
